# Functional Connectivity based Brain Signatures of Behavioral Regulation in Children with ADHD, DCD and ADHD-DCD

**DOI:** 10.1101/2020.04.06.20050013

**Authors:** Christiane S. Rohr, Signe Bray, Deborah Dewey

## Abstract

**Background:** Children with neurodevelopmental disorders such as Attention Deficit Hyperactivity Disorder (ADHD) often struggle with behavioral self-regulation (BR), which is associated with daily-life challenges. ADHD sometimes presents with Developmental Coordination Disorder (DCD), but little is known about BR in DCD. BR is thought to involve limbic, prefrontal, parietal and temporal brain areas. Given the risk for negative outcomes, gaining a better understanding of the brain mechanisms underlying BR in children with ADHD and/or DCD is imperative.

**Methods:** Resting-state fMRI data collected from 115 children (31 typically developing (TD), 35 ADHD, 21 DCD, 28 ADHD-DCD) aged 7-17 years were preprocessed and motion was mitigated using ICA-AROMA. Emotion control, inhibition, and shifting were assessed as subdomains of BR. Functional connectivity (FC) maps were computed for ten limbic, prefrontal, parietal and temporal regions of interest and were investigated for associations with BR subdomains across all participants as well as for significant group differences.

**Results:** Multiple FC patterns were associated with BR across all participants. Some FC patterns were associated with multiple BR subdomains, while others were associated with only one. Differences in BR were found only between children with ADHD (i.e. ADHD and ADHD-DCD) and those without ADHD (i.e. TD and DCD). FC differences were also found between children with and without ADHD.

**Conclusions:** Our results show dimensional associations between BR subdomain scores and whole-brain FC and highlight the potential of these associative patterns as brain-based signatures of BR in children with and without ADHD.

## 1 INTRODUCTION

Many children with neurodevelopmental disorders (NDDs) such as Attention Deficit Hyperactivity Disorder (ADHD) experience difficulty in regulating their emotional behaviors (1, 2). Children with NDDs may be sensitive to external affective cues, making it hard for them to ignore distractions and follow instructions given by teachers or parents (3-5). They may also display frequent and intense shifts in emotions, and have trouble recovering from negative events (3). This struggle with behavioral self-regulation (BR) not only impacts children’s social relationships and performance at school, but also results in greater daily-life and mental health challenges overall (6, 7). Evidence of treatment success with medication is limited (8), and many clinical trials have failed to address BR difficulties in children with ADHD (1, 2). Yet, up to 50% may have difficulty in regulating their emotions and display high levels of emotional lability (9-11).

A subset of children with ADHD also present with Developmental Coordination Disorder (DCD), a NDD that is characterized by impaired motor coordination that significantly interferes with activities of daily living and impacts school performance and activities, as well as leisure and play (12). While very little research has been conducted into BR in DCD to date, a handful of recent studies indicate that problems with BR may exist for children with DCD as well (13-16).

BR can be broken down into inhibitory control, cognitive flexibility and emotion control processes. Inhibitory control is the ability to suppress interfering distractions and prepotent motor responses (4, 17, 18). Cognitive flexibility, which is often measured using set-shifting, is the readiness with which one can switch from one task or mindset to another (4, 19, 20). Finally, emotion control is the process by which we influence the emotions we experience, when we experience them, and how we experience and express them (21, 22).

The neural substrates of BR have been extensively studied in neurotypical adults (23) and adults with affective disorders (24), but less is known about the neural expression of BR subdomains in children and children with NDDs (25, 26). The neural signatures of ADHD as a whole have been elusive due to heterogeneity in the condition, suggesting that looking across the spectrum and range of expression of specific features may be more promising (27-29). Task-based fMRI studies of BR have traditionally focused on areas in the prefrontal cortex (PFC), including orbitofrontal cortex (OFC) and the anterior cingulate cortex (ACC), as well as limbic areas such as the amygdala and nucleus accumbens (NAcc), but more recent reviews and frameworks also implicate regions in parietal and temporal cortex such as the inferior parietal lobule (IPL) and temporal pole (TP) (21, 30, 31). Considering the widespread repercussions suboptimal BR can have throughout childhood and into adulthood, characterization of the interactions between these areas within the brain or their whole-brain functional connectivity (FC) has enormous potential for diagnosis and individually tailored treatment of BR difficulties in NDDs (1).

Distributed FC patterns (i.e. patterns spanning across much of the brain) have previously been associated with the core ADHD symptoms of inattention and hyperactivity (32, 33), but less is known about distributed FC in relation to BR and other symptomatic behaviors that undergo maturation in NDDs. In particular, comprehensive whole-brain FC signatures (i.e. conglomerates of distributed FC patterns) that define aspects of BR, are not well studied, despite the widespread repercussions of suboptimal BR for children that persist into adulthood, and the enormous potential of whole-brain FC profiles as ‘neuromarkers’ for diagnosis and individually tailored treatment.

In this study, we therefore sought to provide a more comprehensive picture into whole-brain FC signatures associated with different subdomains of BR. We choose seed regions of interest that involve multiple brain networks in order to assess convergence or divergence of neural pathways in relation to BR subdomains. To do so, we investigated associations between BR scores on the Behavior Rating Inventory of Executive Function (BRIEF), a parent report of BR, and the FC of ten limbic, prefrontal, parietal and temporal regions of interest (ROIs) in resting-state fMRI data of typically developing (TD) children and children with ADHD, DCD or ADHD-DCD. We hypothesized that children with an NDD would evidence more problems in BR than TD children. We further hypothesized that the FC of selected limbic, prefrontal, parietal and temporal ROIs would show associations with BR. Specifically, we hypothesized that (a) associations between FC and BR across all participants would exist; and (b) reflecting common neural mechanisms, some FC patterns would be associated with more than one BR subdomain, whereas others would not. We further hypothesized that (c) some FC patterns would differentiate children with and without a diagnosis and (d) some of these FC patterns would associate with BR, whereas others would not.

## 2 METHODS AND MATERIALS

This research was conducted in accordance with the Declaration of Helsinki for experiments involving human subjects. It was approved by the Conjoint Health Research Ethics Board of the University of Calgary. Written consent and verbal assent were obtained from parents or guardians, and participants, respectively.

### 2.1 Participants

Participants were recruited from local schools and through community advertisements in locations such as hospitals and physician’s offices in Calgary, Alberta, Canada. TD children and children diagnosed with ADHD, DCD or ADHD-DCD, as well as children with attention and/or motor difficulties, were eligible, provided they had not been diagnosed with another neurodevelopmental or psychiatric disorder, a neurological, metabolic or genetic condition, and were not born preterm (<36 weeks) or with very low birth weight (<1500g). Potential participants were screened for contraindications for MRI and other medical problems that would prevent participation. Recruited participants who met the above criteria were invited to participate in a detailed neuropsychological assessment. Children were classified as DCD if, in keeping with the Diagnostic and Statistical Manual of Mental Disorders - Fourth Edition Text Revision (DSM-IV-TR) (34), they scored less than the 16th percentile on the Movement Assessment Battery for Children - Second Edition (MABC-II) (35), and were reported by their parents as exhibiting motor difficulties that interfered significantly with daily functioning on the Developmental Coordination Questionnaire (36). Children were classified as ADHD if they met the diagnostic criteria on the Diagnostic Interview for Children and Adolescents - IV (37), or had a T score above the 95th percentile on the Conners’ Parent Rating Scale - Revised (CPRS-R) (38) and were diagnosed by a physician as having ADHD based on DSM-IV-TR criteria. Children meeting criteria for both ADHD and DCD were classified as ADHD-DCD. Children who were prescribed stimulant treatment for ADHD were asked to refrain from taking their medication on the day they underwent MRI scanning. A total of 149 participants who met criteria underwent resting state fMRI. Of these, 6 did not complete the diagnostic assessment measures; 1 (ADHD-DCD) had a diagnosis of Autism Spectrum Disorder; 14 (6 TD, 3 DCD, 3 ADHD, 2 ADHD-DCD) did not complete the cognitive assessment; and 4 (1 ADHD-DCD, 1 ADHD, 2 DCD) did not complete their MRI scan. Of the remaining participants, 9 had excessive head motion on their fMRI scan (>5mm maximum absolute displacement). Participants’ data were further evaluated for outliers on behavioral measures, defined as > 3 SD from the mean. No participant was excluded due to this criterion. The final sample consisted of 115 participants; characteristics are provided in Table 1.

**Table 1.** Participant characteristics. Means and standard deviations (in brackets) are provided for the total sample, as well as for TD participants and participants with DCD, ADHD and ADHD-DCD and the children without and with ADHD, separately. Motion (mm) refers to the absolute maximum displacement at any timepoint in the resting-state fMRI scan prior to motion mitigation and denoising procedures. N=number of participants; IQ=Intelligence Quotient. Inhibition, Shifting and Emotion Control scores on BRIEF are given as T scores. † denotes a significant difference between children with and without ADHD

|  | Total | TD | DCD | ADHD | ADHD-DCD | without ADHD | with ADHD |
| --- | --- | --- | --- | --- | --- | --- | --- |
| N (females) | 115 (39†) | 31 (14) | 21 (13) | 35 (8) | 28 (4) | 52 (27) | 63 (12) |
| Age in years | 11.2 (2.6) | 11.6 (3) | 11.7 (2.7) | 11.4 (2.5) | 10.3 (1.9) | 11.6 (2.9) | 10.9 (2.3) |
| Left-handed | 16 | 3 | 4 | 4 | 5 | 7 | 9 |
| Motion (mm) | 1.5 (1.2) | 1.5 (1.1) | 1.2 (0.9) | 1.5 (1.1) | 1.8 (1.5) | 1.36 (1.04) | 1.66 (1.3) |
| FIQ | 107 (15) | 111 (15) | 111 (16) | 108 (12) | 100 (17) | 111 (15) | 105 (14.5) |
| Inhibition | 55.2 (13.6)† | 48.9 (10.3) | 51.1 (12.7) | 60.8 (12.2) | 57.9 (15.2) | 49.8 (11.3) | 59.6 (13.8) |
| Shifting | 57 (14.9)† | 51.9 (13.2) | 50.2 (11.4) | 60.5 (15.6) | 63.3 (14.7) | 51.2 (12.4) | 61.8 (15.1) |
| Emotion Control | 56.6 (13.7)† | 53 (13) | 49.6 (9.7) | 58.9 (14.9) | 63 (12.4) | 51.6 (11.8) | 60.7 (13.9) |

### 2.2 Cognitive and Behavioral Assessment

The cognitive and behavioral data used here included Full Scale Intelligence (FSIQ), handedness and BR. FSIQ was measured using the Wechsler Abbreviated Scale of Intelligence (WASI) (39). Handedness was determined based on the preferred hand identified and used by the child when performing fine motor tasks on standardized measures of motor function (i.e., Movement Assessment Battery for Children - Second Edition) (35). BR was assessed with the Behavior Rating Inventory of Executive Function (BRIEF) (40), a standardized parent report measure of executive function behaviors for children aged 5-18 years. BRIEF subscales provide measures of three subdomains of BR, which are labelled “inhibit”, “shift”, and “emotion control”. The “inhibit” subscale assesses the ability to resist impulses and to stop one’s own behavior” (sample item: “acts wilder or sillier than others in groups (birthday parties, recess)”). The “shift” subscale assesses the ability to move freely from one situation, activity, or problem to another; to tolerate change, and to switch or alternate attention (sample item: “resists or has trouble accepting a different way to solve a problem with schoolwork, friends, chores, etc.”). Finally, the “emotion control” subscale assesses the ability to regulate emotional responses appropriately (sample item: “overreacts to small problems”). Normed T-scores for the BRIEF were used in the analyses, with higher scores indicating more problems in executive functions.

### 2.3 MRI Data Acquisition Parameters

Data were acquired at the Seaman Family MR Research Centre at the University of Calgary across two MRI systems due to a system upgrade. 67 scans were collected on a 3T GE Signa VH/i (Waukesha, WI) with an eight-channel phased-array radiofrequency head coil and 48 scans were collected on a GE 750 with an eight-channel phased-array head coil. Children were instructed to keep their eyes on a fixation cross at the center of the screen. Functional images were acquired using a gradient-echo EPI sequence in 40 axial slices (120 volumes, TR=2500 ms, TE=30 ms, FA=70, matrix size 64×64, voxel size 3.44×3.44×3mm^3^; duration: 5 minutes) in the first round of acquisition, and in 26 axial slices (140 volumes, TR=2500ms, TE=30ms, FA=70, matrix size 64×64, voxel size 3.44×3.44×4mm^3^; duration: 5.8 minutes) in the second round of acquisition. Anatomical scans were acquired using a T1-weighted MPRAGE sequence (TR=1000ms, TE=2.5ms, FA=18, voxel size 0.9×0.9×4mm^3^ in the first round of acquisition and TR=7.4ms, TE=3.1ms, FA=13, voxel size 1×1×0.8mm^3^ in the second round of acquisition).

### 2.4 MRI Data Preprocessing

Data preprocessing used functions from FSL (41) and AFNI (42); the specific functions are denoted in brackets. Anatomical data was deobliqued (3drefit), oriented into FSL space (RPI) (3dresample) and skull-stripped (3dSkullStrip and 3dcalc). Functional data was also first deobliqued (3drefit) and oriented into FSL space (RPI) (3dresample). The pipeline further consisted of motion correction (MCFLIRT), skull-stripping (3dAutomask and 3dcalc), spatial smoothing (6mm Gaussian kernel full-width at half-maximum) (fslmaths), grand-mean scaling (fslmaths), registration to the participants anatomical (FLIRT), and normalization to the McConnell Brain Imaging Center NIHPD asymmetrical (natural) pediatric template optimized for ages 4.5 to 18.5 years (43) (FLIRT), followed by normalization to 2×2×2 mm MNI152 standard space (FLIRT). A four-step process previously employed in a study of children with an NDD (25) and supported by a benchmarking methods study (44) was used to address motion and physiological confounds in the data.

### Head-motion and Physiological Confound Mitigation Procedure

A four-step process was used to address motion and physiological confounds in the data. First, motion estimates derived from the preprocessing were utilized to exclude participants with excessive head motion; scans were excluded if they exhibited >5 mm absolute maximum displacement. Second, AROMA was employed, an ICA-based cleaning method (91), which has recently been shown to be most effective in mitigating the impact of head motion, and allows for the retention of the remaining ‘true’ neural signal within an affected volume (96). AROMA is an automated procedure that uses a small but robust set of theoretically motivated temporal and spatial features (timeseries and power spectrum) to distinguish between ‘real’ neural signals and motion artifacts. We chose a threshold that is conservative about what is retained (‘aggressive’) in order to decrease the chance of false positives. Noise components identified by AROMA were removed from the data. Third, images were de-noised by regressing out the six motion parameters, as well as signal from white matter, cerebral spinal fluid and the global signal, as well as their first-order derivatives (44). While there is currently no gold standard (97) regarding the removal of the global signal, it was removed here based on evidence that it relates strongly to respiratory and other motion-induced signals, which persist through common denoising approaches including ICA and models that approximate respiratory variance (98). Motion (defined as each participant’s absolute maximum displacement) was substantially reduced following this procedure (before: 1.5mm ± 1.2mm; after: 0.07mm ± 0.03mm). As a final step, described below, head motion, defined as absolute maximum displacement, was included in the analysis models as a covariate of no interest, in order to minimize a residual influence of motion on the results.

### 2.6 Analysis of Demographic, Cognitive and Behavioral Measures

ANOVAs were utilized to assess differences in demographics and head motion between the four participant groups: TD children and children with DCD, ADHD or ADHD-DCD. ANCOVAs were then used to assess differences in BR, controlling for any observed differences in demographics between children as covariates of no interest. As described in the Results section below, there were no differences in BR between TD children and children with DCD, and also no differences between children with ADHD and ADHD-DCD; therefore, we focused the analysis on children with ADHD (ADHD and ADHD-DCD) versus children without ADHD (TD children and children with DCD). T-tests were utilized to assess differences in demographics, head motion and BR between these groups. Finally, Pearson correlations were computed to assess the relationship between demographics, head motion and BR scores. These analyses were carried out using SPSS 22 (Chicago, IL).

### 2.7 Analysis of fMRI Data

To examine FC of the regions of interest (ROIs), how they associate with BR scores across the brain and how they differ between groups, ten ROIs were selected based on a well-known model of BR (21) (see Figure 1 for details). To compute each ROI’s FC map, the average time course was extracted for each ROI and entered into a voxel-wise correlation with every other voxel in the brain. Resultant whole-brain FC maps were normalized using Fisher’s r-to-z transform (z=.5[ln(1+r)-ln(1-r)]) for comparison across individuals. Group-level statistical testing was conducted using FLAME 1, a mixed-effects analysis in FSL’s FEAT. In a correlation analysis, each BR subscale T-score was converted to a z-score and entered into a separate model to assess the association between FC and the respective subscale across the group. In a group contrast analysis, FC maps were compared between groups that showed significant differences in BR scores. All models included z-scored sex, scanner and motion as nuisance covariates. Voxel-wise thresholding was set at z-score >2.3, and cluster correction was conducted using Gaussian Random Field theory with p <0.05. The p-values for these results were then Bonferroni-corrected for ten comparisons (i.e., the number of seeds that were examined; significance set at p <0.005).

**Figure 1.**
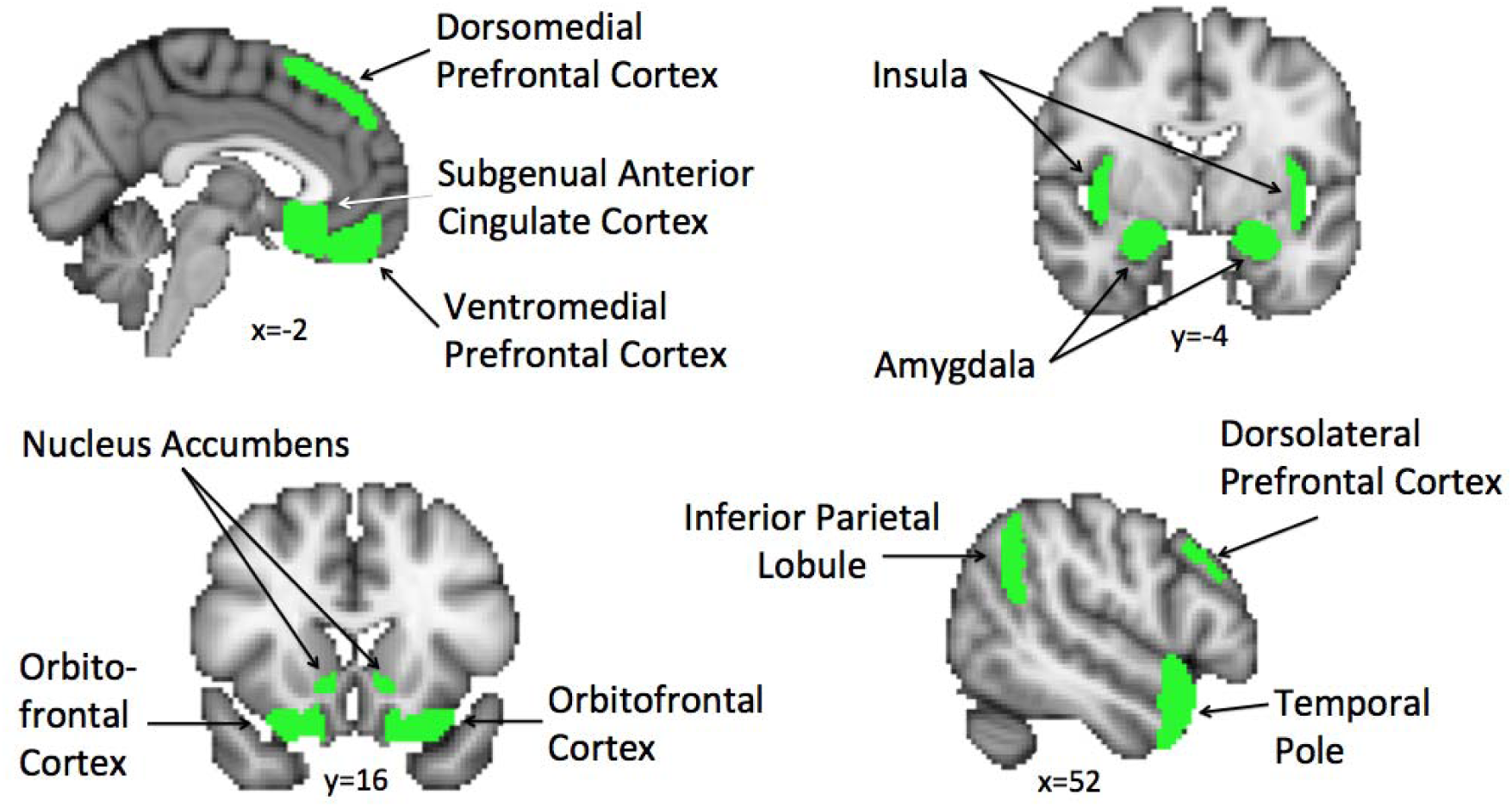
Seed Regions of Interest. To examine how FC associates with BR scores across the brain and how FC differs between groups, ten ROIs were selected in limbic areas (i.e., amygdala, insula and nucleus accumbens), prefrontal areas (i.e., dorsolateral, dorsomedial and ventromedial prefrontal cortex; orbitofrontal cortex and subgenual anterior cingulate cortex), parietal areas (i.e., inferior parietal lobule) and temporal areas (i.e., temporal pole). ROI masks were anatomically defined using probabilistic parcellation units provided through FSL with the Harvard-Oxford Atlas and thresholded at 50% probability, meaning any given voxel within the seed mask had a >50% probability of lying within the specified region.

### 2.8 Assessment of Specificity to Behavioral Regulation

To evaluate whether our correlation analyses captured BR dimensionally or were driven by the categorical difference in scores due to ADHD diagnoses, we performed a post-hoc correlation analysis accounting for diagnostic status through an added nuisance covariate.

## 3 RESULTS

### 3.1 Sample Characteristics

Characteristics for the sample are provided in Table 1. There were no significant differences in BR between TD children and children with DCD, or between children with ADHD and children with ADHD-DCD. Group comparisons were therefore carried out only on the combined groups of children with ADHD (ADHD and ADHD-DCD, n = 63) versus children without ADHD (TD and DCD, n = 52). Significant differences between children with and without ADHD existed in sex (p=0.0002) and the distribution across scanners (p=0.006), but not in motion (neither before cleaning nor after; both p>.11). Adjusting for these covariates (i.e. sex and distribution across scanners), results still showed significant differences between children with and without ADHD in inhibition (p=0.004), shifting (p=0.002), and emotion control (p=0.007), reflecting greater challenges with BR for children with ADHD. Across all participants, BR subdomains correlated highly with each other (inhibition-shifting: r=.53, p<0.0001; inhibition-emotion control: r=.52, p<0.0001; shifting-emotion control: r=.75, p<0.0001). No correlations were observed between BR, and age, IQ or motion (neither before cleaning nor after; all p>.23).

### 3.2 Functional Connectivity

A total of 23 FC patterns across 8 seeds were associated with BR across the entire group (i.e. all participants) (Table 2) in the correlation analysis. 61% (n = 14) of these FC patterns were associated with two or three BR subdomains (Figure 2) while 39% (n = 9) showed a unique association with just one BR subdomain (Figure 3). Overall, greater BR problems tended to be associated with stronger negative FC, but associated with weaker positive FC in 30% (n = 7) of the BR-associated patterns. These seven overlapped with FC patterns that were significantly different between children with and without ADHD. All FC patterns detected in the correlation analysis remained associated with BR after controlling for ADHD diagnosis, suggesting that these were dimensional effects and not driven by diagnostic status.

**Table 2.** Associations between BR subdomain scores and FC across all participants. BIL=bilateral; dlPFC=dorsolateral prefrontal cortex; dmPFC=dorsomedial prefrontal cortex; L=left; Lat=Laterality; OFC=orbitofrontal cortex; R=right; rACC=rostral anterior cingulate cortex; ROI=region of interest; sgACC=subgenual anterior cingulate cortex; V1-V2= visual cortex 1-2; V2-V3=visual cortex 2-3; vACC=ventral anterior cingulate cortex; vlPFC=ventrolateral prefrontal cortex; vmPFC=ventromedial prefrontal cortex; TP=temporal pole. Direction refers to the direction of the association with BR. *denotes overlap with FC pattern observed in group difference analysis

|  | Lat | Seed ROI | Direction | Voxels | p-value | Z-Max | X | Y | Z | Lat | Connectivity |
| --- | --- | --- | --- | --- | --- | --- | --- | --- | --- | --- | --- |
| *Emotion control* | L | sgACC * | neg | 999 | 0.000936 | 3.84 | -48 | -10 | -10 | L | STG, Insula, Pallidum |
|  | L | Insula | neg | 1235 | 0.000919 | 3.43 | 20 | -64 | -12 | BIL | V2-V3 |
|  | L | Accumbens | neg | 2108 | 1.06E-05 | 3.72 | -14 | -72 | 8 | BIL | V1-V2 |
|  | R | TP | pos | 1834 | 0.000103 | 4 | 30 | 58 | -2 | BIL | rACC, Frontal Pole |
|  | R | sgACC * | neg | 1242 | 0.000255 | 3.62 | -2 | 14 | -2 | L | sgACC, Pallidum, Putamen, Insula |
| *Inhibition* | L | OFC * | neg | 2034 | 1.08E-05 | 3.93 | 12 | 34 | 6 | BIL | vmPFC, Accumbens |
|  | L | vmPFC | neg | 1258 | 0.00138 | 3.51 | -42 | 10 | 26 | L | vlPFC, Insula |
|  | L | TP | pos | 2169 | 0.000122 | 3.76 | 8 | -72 | 30 | BIL | Precuneus, Cuneus, V1 |
|  | L | Accumbens | neg | 2898 | 1.73E-06 | 3.82 | -12 | -72 | 14 | BIL | V1-V2 |
|  | R | dlPFC | neg | 992 | 0.00324 | 3.14 | 14 | -74 | 22 | BIL | V1-V2 |
|  | R | dmPFC | neg | 1180 | 0.000286 | 3.39 | -12 | -68 | 8 | R | V1-V2 |
|  | R | OFC * | neg | 1353 | 0.00112 | 3.91 | -30 | 26 | -22 | L | vmPFC, Accumbens |
|  | R | vmPFC | neg | 1967 | 7.25E-05 | 3.78 | -36 | 38 | 0 | L | vlPFC, Insula |
|  | R | Accumbens | neg | 1357 | 0.000747 | 4.27 | -38 | 6 | 14 | L | Insula, TP |
| *Shifting* | L | vmPFC | neg | 2363 | 4.65E-06 | 4.24 | -18 | 2 | 8 | L | vlPFC, Insula |
|  | L | sgACC * | neg | 1759 | 5.84E-05 | 3.65 | -18 | 10 | 10 | L | Putamen, Pallidum, Insula |
|  | L | sgACC * | neg | 1076 | 0.00276 | 3.62 | 36 | -12 | -8 | R | Putamen, Pallidum, Insula |
|  | L | Accumbens | neg | 2544 | 1.13E-06 | 3.78 | -26 | -72 | 4 | BIL | V1-V2 |
|  | L | Accumbens | neg | 1442 | 0.000302 | 3.46 | -50 | -6 | 42 | L | Caudate, Putamen, Insula, TP, vlPFC |
|  | R | vmPFC | neg | 3104 | 1.07E-06 | 4.3 | -42 | 10 | 8 | L | vlPFC, Insula |
|  | R | TP | pos | 2163 | 5.26E-05 | 3.74 | 22 | 58 | 0 | BIL | rACC, Frontal Pole |
|  | R | sgACC * | neg | 994 | 0.00285 | 3.58 | 42 | -8 | -4 | R | Putamen, Insula |
|  | R | Accumbens | neg | 1756 | 4.78E-05 | 3.83 | 8 | -12 | -6 | BIL | Thalamus, Pallidum, Putamen, Insula |

**Figure 2.**
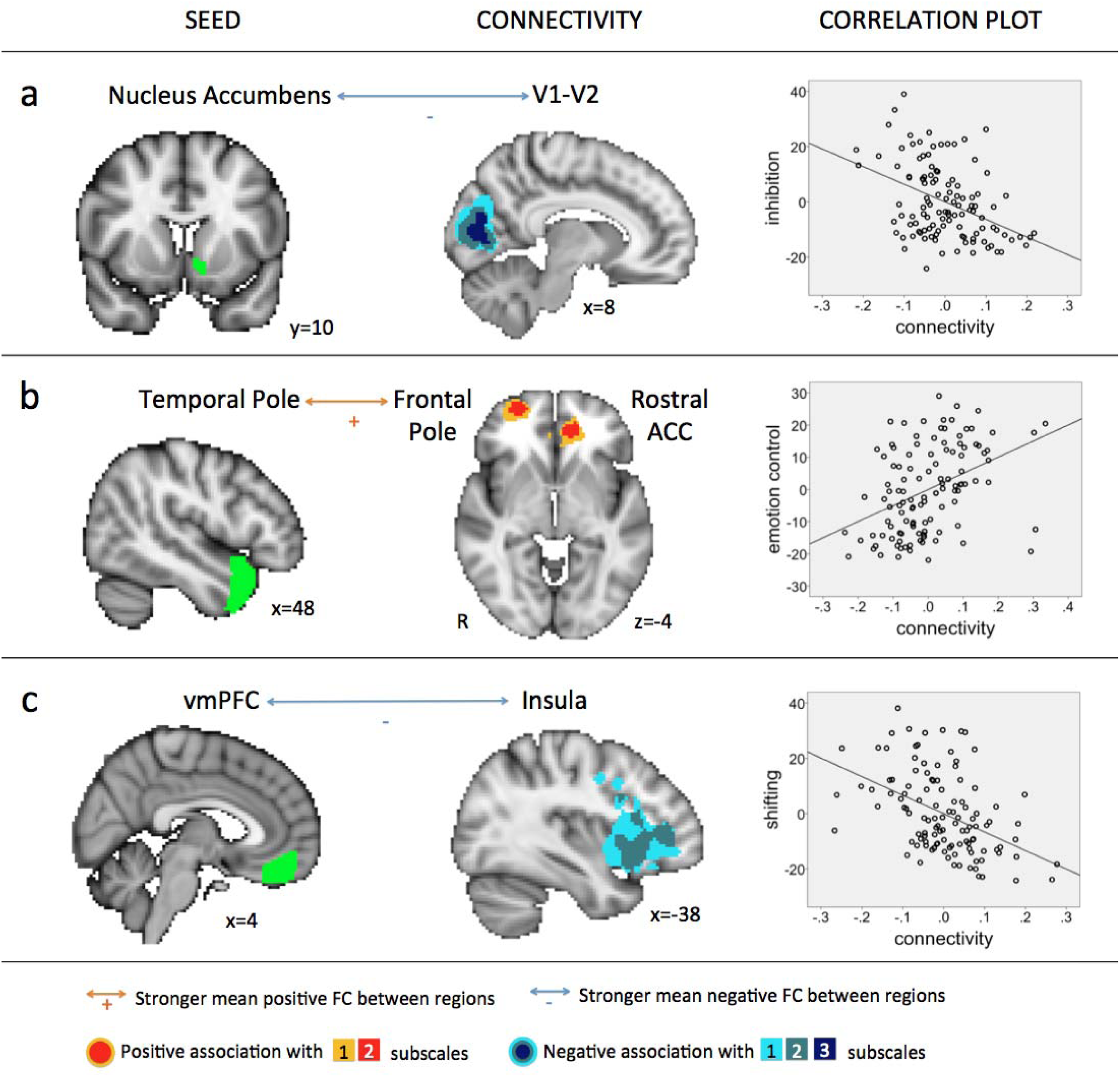
FC patterns that associated with more than one BR subscale across all participants. Stronger negative FC was associated with greater BR problems in A and C, whereas stronger positive FC was associated with greater BR problems in B. Panel A shows FC associated with all three subscales (the correlation with inhibition is shown as an example); panel B shows FC associated with shifting and emotion control (the correlation with emotion control is shown); and panel C shows FC associated with inhibition and shifting (the correlation with shifting is shown). Note that these patterns were present bilaterally; the right side is shown. Positive associations between FC and scores are depicted in red-yellow, negative associations between FC and scores are depicted in blue-light blue. Maps are binarized to visualize overlapping patterns. Colored arrows with + and – signs indicate the direction of FC between seed and cluster regions as identified via the seed’s FC map. Correlation plots show all values adjusted for sex, scanner and motion. ACC=anterior cingulate cortex; R=right hemisphere; V1-V2= visual cortex 1-2; vmPFC=ventromedial prefrontal cortex

**Figure 3.**
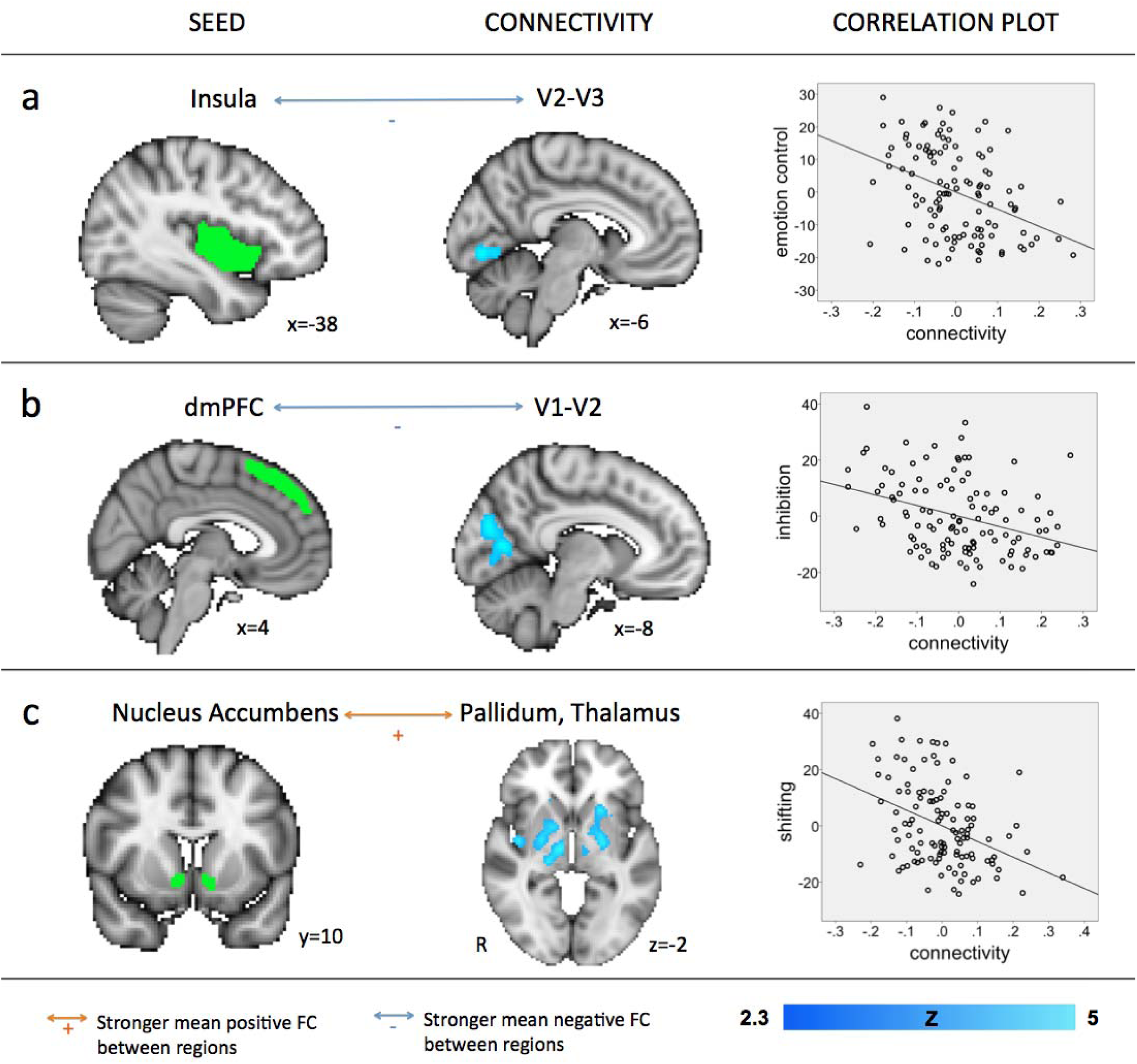
FC patterns that associated with only one BR subscale across all participants. Stronger negative FC was associated with greater BR problems in A and B; weaker positive FC was associated with greater BR problems in C. Panel A shows FC associated with emotion control; panel B shows FC associated with inhibition; and panel C shows FC associated with shifting. Note that these patterns were present bilaterally; the right side is shown. Negative associations between FC and scores are depicted in blue-light blue; Z values reflect the strength of the association. Colored arrows with + and – signs indicate the direction of FC between seed and cluster regions as identified via the seed’s FC map. Correlation plots show all values adjusted for sex, scanner and motion. dmPFC=dorsomedial prefrontal cortex; R=right hemisphere; V1-V2= visual cortex 1-2; V2-V3=visual cortex 2-3

Eleven FC patterns were significantly different between children with and without ADHD (Table 3) in the group contrast analysis. 36% (n = 4) of these overlapped with FC patterns that associated with BR in the correlation analysis. Children with ADHD exhibited significantly weaker positive FC revolving around sgACC and putamen/pallidum/insula, which was associated with shifting and emotion control, as well as around OFC and vmPFC, which was associated with inhibition (Figure 4; FC values were drawn from the two independent analyses).

**Table 3.** Differences in FC between children with and without ADHD. A=children with ADHD; nonA=children without ADHD; BIL=bilateral; dlPFC=dorsolateral prefrontal cortex; dmPFC=dorsomedial prefrontal cortex; L=left; Lat=Laterality; M1=primary motor cortex; OFC=orbitofrontal cortex; PCC=Posterior Cingulate Cortex; R=right; rACC=rostral anterior cingulate cortex; ROI=region of interest;S1=primary somatosensory cortex; sgACC=subgenual anterior cingulate cortex; V1-V2= visual cortex 1-2; V2-V3=visual cortex 2-3; vACC=ventral anterior cingulate cortex; vmPFC=ventromedial prefrontal cortex. Direction refers to the direction of the association with BR. *denotes overlap with FC patterns associated with BR subdomain scores across all participants

|  | Lat | Seed ROI | Direction | Voxels | p-value | Z-Max | X | Y | Z | Lat | Connectivity |
| --- | --- | --- | --- | --- | --- | --- | --- | --- | --- | --- | --- |
| *differences between children with and without ADHD* | L | OFC * | A < nonA | 2263 | 1.67E-06 | 3.68 | 16 | 44 | -10 | R | vmPFC, Accumbens |
|  | L | vmPFC | A < nonA | 2845 | 1.79E-07 | 4.06 | 16 | 22 | -6 | BIL | Accumbens, Caudate, OFC |
|  | L | sgACC * | A < nonA | 1789 | 5.20E-05 | 4.34 | 44 | -12 | 12 | R | Putamen, Pallidum, Insula |
|  | L | Accumbens | A < nonA | 2073 | 5.96E-07 | 4.02 | 30 | 32 | -24 | BIL | OFC, sgACC, rACC, vmPFC |
|  | R | OFC * | A < nonA | 2876 | 6.56E-07 | 4.78 | -30 | 28 | -26 | BIL | vmPFC, Accumbens |
|  | R | vmPFC | A < nonA | 2566 | 4.77E-07 | 3.95 | 16 | 20 | -6 | BIL | Accumbens, Caudate, OFC |
|  | R | sgACC * | A < nonA | 1781 | 0.000112 | 4.19 | 44 | -12 | 12 | R | Putamen, Pallidum, Insula, Thalamus, Amygdala/Hippocampus |
|  | R | Accumbens | A < nonA | 3061 | 1.19E-07 | 4.08 | -22 | 34 | -24 | BIL | Caudate, Putamen, vmPFC |
|  | R | Amygdala | A < nonA | 1013 | 0.00261 | 3.57 | 32 | 48 | -10 | BIL | sgACC |
|  | L | OFC | A > nonA | 1131 | 0.00108 | 3.85 | -10 | -46 | 14 | BIL | PCC |
|  | R | Insula | A > nonA | 975 | 0.00464 | 3.4 | -52 | -74 | 18 | L | Superior Lateral Occipital Cortex |

**Figure 4.**
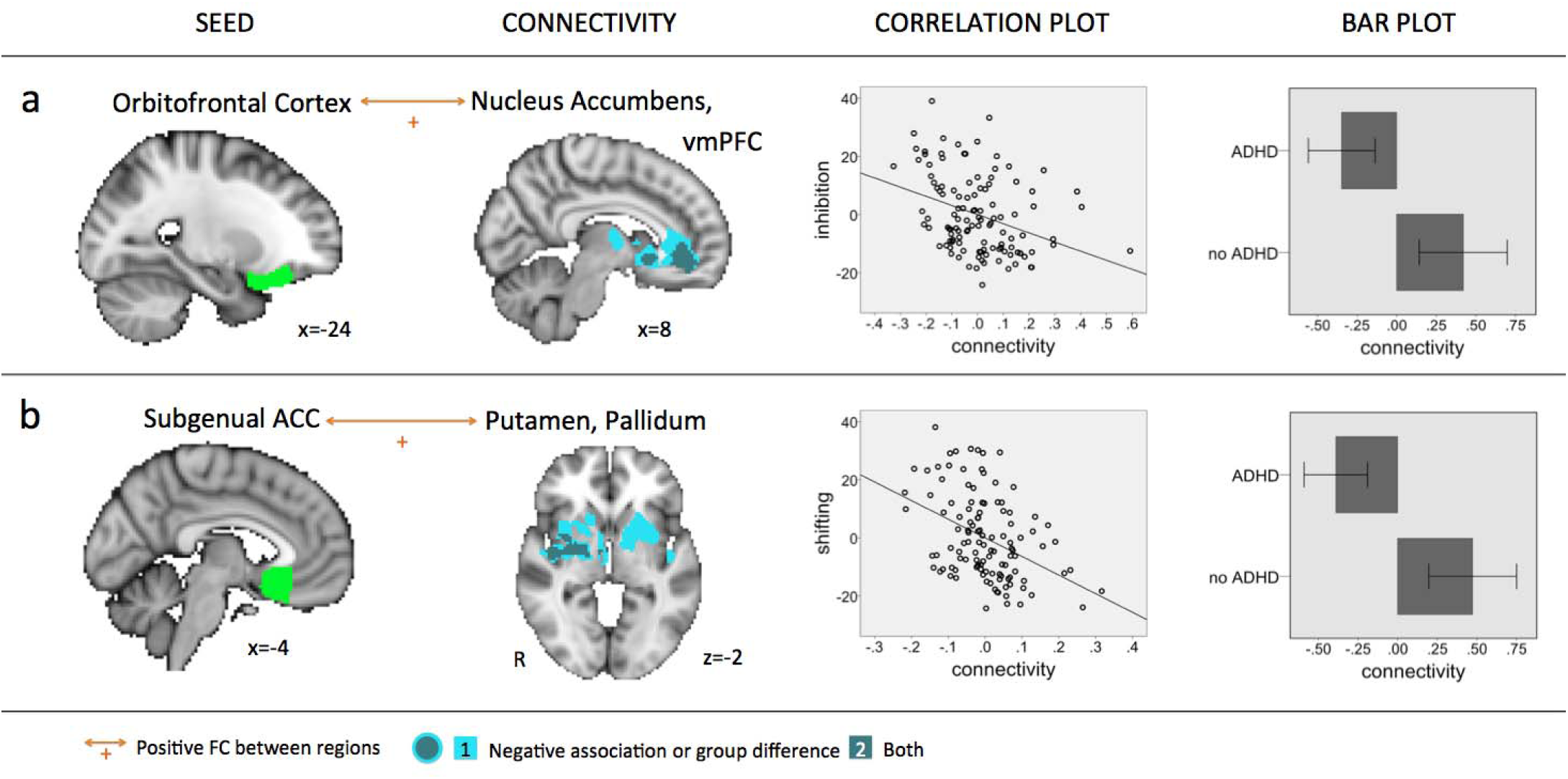
FC patterns that associated with BR subscales and showed significantly weaker positive FC in children with ADHD. Panel A shows FC that was associated with inhibition and significantly different between children with and without ADHD. This pattern was present bilaterally; the left side is shown. Panel B shows FC that was associated with both shifting and significantly different between children with and without ADHD. This pattern was also present bilaterally; again the left side is shown. A contralateral version of this pattern – from the left sgACC to the left rather than the right putamen, pallidum and insula - was associated with emotion control. In both A and B, weaker positive FC was associated with greater BR problems. Negative associations between FC and BR scores and significant group differences are depicted in blue-lightblue; maps are binarized to visualize overlapping patterns. Colored arrows with + signs indicate positive FC between seed and cluster regions as identified via the seed’s FC map. Bar plot and correlation plot FC values shown are from two independent analyses. Bar plot FC values were extracted from the group contrast GLM and show significant differences in average FC within the overlapping cluster with error bars (CI 95%). Correlation plot FC values were extracted from the correlation GLMs testing for significant associations with BR and show FC values within the overlapping cluster. All values are adjusted for sex, scanner and motion. ACC=anterior cingulate cortex; R=right hemisphere; vmPFC=ventromedial prefrontal cortex

## 4 DISCUSSION

Poorer behavioral self-regulation (BR) is a known issue for children with NDDs such as ADHD and is associated with greater daily-life challenges and an increased risk for psychiatric comorbidities (6, 7). In this study, we found that children with a diagnosis of ADHD (i.e. children with ADHD or ADHD-DCD) had significantly more problems in BR than typically developing children and children with ‘pure’ DCD. The strength of distributed patterns of FC was associated with children’s individual differences in three BR subdomains: emotion control, inhibition and shifting. Some FC patterns were associated with more than one BR subdomain while other FC patterns were specific to one BR subdomain; none were explained by ADHD diagnostic status. A subset of FC patterns that were significantly different between children with and without ADHD converged with FC patterns that associated with BR in the group overall. Overall, our results show that selected subsets of whole-brain FC data involving frontostriatal, temporo-limbic and visual pathways may have utility as brain-based signatures of BR across children with and without ADHD.

Unlike several recent studies (13-16), we found no elevation of BR scores in children with ‘pure’ DCD. This may be because we rigorously screened for comorbid ADHD; it has been found that up to 50% of children with DCD meet diagnostic criteria for ADHD while only 5% are diagnosed (45). We also found that children with ADHD-DCD showed elevated scores on BR; therefore, it is likely that BR problems in children with DCD are due to comorbidity with ADHD rather than DCD itself.

FC patterns that generalized to more than one BR subdomain included pathways between prefrontal and temporal areas, and pathways between striatal and visual areas. FC between vmPFC and vlPFC and insula, regions with strong anatomical connections, was related to shifting and inhibition. Data suggests that the vmPFC integrates affective valuations from temporal lobe systems including the insula that provide interoceptive awareness of the body and information about prior encounters with stimuli (21, 46). FC between TP and rACC (also known as perigenual ACC), also regions that are anatomically connected, was related to shifting and emotion. Macaque studies have shown that the ACC receives input from the TP and other temporal areas (47, 48), and both are thought to act as domain-general hubs, with the TP integrating socioemotional information (49) and the ACC monitoring for emotionally conflicting information and prepotent responses (50-52). Thus, both FC pathways involve prefrontal cognitive control areas (vmPFC, rACC) that receive pre-processed sensory information from temporal lobe systems (insula, temporal pole).

FC between nucleus accumbens (NAcc) and visual areas, which was linked to all three BR subdomains, has been observed during reward processing (53) and NAcc and visual areas have been jointly activated in reward-directed action and inhibition of action (54) and response to incentives (55). NAcc is known to play an important role in processing rewarding and reinforcing stimuli (e.g., food and water) (56) as well as in reward anticipation (57) and outcome prediction (58, 59); it receives projections from dopamine-releasing neurons (60) and interacts with the PFC (61). Reward and BR are arguably linked (62), especially in children (63). This pattern is a FC pathway between a primary sensory system and a dopaminergic structure that exchanges information with prefrontal control areas.

FC patterns that were specific to just one BR subdomain included pathways between prefrontal and visual areas, between temporal and visual areas, and thalamo-striatal pathways. For instance, FC between the insula and the visual cortex was specific to emotion control. This is an anatomically connected pathway relevant to interoception in emotion processing (64) that has been linked to alexythimia (65). Alexithymia is a difficulty in identifying and verbalizing one’s own emotions; it can also impact an individual’s ability to identify emotions in others and empathize; both hamper an individual’s ability to regulate their emotions in an adaptive manner (66). As another example, FC between the dmPFC and visual areas was specific to inhibition. This pathway is heavily implicated in top-down modulation of visual processing and visual recognition especially in relationship to working memory (67, 68) and conflict processing (50), and has been shown to be altered in major depressive disorder (67), another condition in which issues with BR are readily apparent (69). Finally, FC within thalamo-striatal pathways was specific to shifting. This anatomically closely connected pathway is rich in dopamine, and dopamine is thought to code for learned associations and mediate approach behavior toward a reward; it is known to be actively involved in tasks requiring cognitive flexibility (70).

FC patterns that were significantly different between children with and without ADHD and associated with BR centered on frontostriatal reward pathways known to be affected in ADHD (71). Inhibition has been associated with FC between the OFC and NAcc/vmPFC, and animal studies have shown that haemodynamic signals of, and neuronal projections between, OFC and NAcc are related to inhibition-related processes that are part of reinforcement learning (72, 73). Shifting and emotion control have been associated with FC between sgACC and putamen/pallidum and activity in both structures has been found to be aberrant during reward prediction in obsessive-compulsive disorder (74), a disorder often comorbid with ADHD that like ADHD is a ‘disorder of control’ (75). Volume in both structures has been found to be different in individuals with ADHD (76).

It is not clear what drives these FC patterns to be significantly different between children with and without ADHD, when many other FC patterns were not, despite significant differences in scores on measures of BR. This may lead one to consider FC patterns that were not significantly different to be ‘intact’. However, one needs to keep in mind that FC in these patterns were elevated in children with ADHD, which may be meaningful clinically, as statistical significance is not identical to clinical relevance (77).

Individual differences in BR have been repeatedly found to be associated with individual features in FC (50, 78, 79). Taking individual differences into account can help expose the underlying neural substrates of complex cognitive skills, emotions and social competencies, and has proven useful in the investigation of both neurotypical (80-83) and clinical populations (84-86) as traits and abilities associated with NDDs such as ADHD also exist in the neurotypical population, and fall onto a spectrum (84, 87). The use of individual differences also allows for more statistical power in studies on NDDs, which struggle with sample sizes and many sources of heterogeneity (29, 88-90).

The current study has several distinct strengths, which include appropriate preprocessing techniques, the use of a measure that assessed three subdomains of BR in a relatively large group of children with and without ADHD, and the inclusion of a comorbid group that helped to attribute differences to ADHD specifically. The denoising methods employed allowed for the retention of the remaining ‘true’ neural signal within an affected volume and were in accordance with the latest best practices for reducing motion artifacts (44), which is particularly relevant for a neurodevelopmental study (91-93). The measure used to assess BR is well validated (40) and although parent-reports are subjective, they capture a measure of behavior integrated over a longer time frame than can be observed in a laboratory visit and have better test-retest reliability (94). The study also has several weaknesses, including a relatively short scan time and differences between children with and without ADHD in (i) sex ratios and (iii) distribution across scanners, as well as a small N in the ‘pure’ DCD group. We have done our best to account for these by including sex and scanner as covariates in all analyses. While a short scan time is of benefit from an acquisition perspective, longer scan times may strengthen the reliability of FC estimates (95).

Our findings present a substantial addition to our knowledge on BR and its underlying neural expression across a neurodiverse spectrum of children with and without ADHD, including children with DCD and combined ADHD-DCD. They suggest that BR problems in DCD are likely attributable to a comorbidity with ADHD. Children’s individual differences in the three BR subdomains further associated with FC. Primary visual to mesolimbic FC pathways and FC pathways that integrate pre-processed sensory information with cognitive control processes were associated with multiple BR subdomains while thalamo-striatal FC patterns and FC patterns involving the visual stream were specific to one subdomain. Only BR-associated FC in frontostriatal reward pathways was significantly different between children with and without ADHD. Overall, our results highlight the utility of directly examining variables of interest and their associations with whole-brain FC and show that these may have utility as brain-based signatures of BR across children with and without ADHD.

## Data Availability

Data are not currently publicly available.

## ACKNOWLEDGEMENTS

This work was supported by a grant from the Canadian Institutes of Health Research awarded to DD (MOP 88588), as well as NSERC CREATE I3T and Alberta Innovates Postdoctoral Fellowships awarded to CR. The funders had no role in study design, data collection and analysis, decision to publish or preparation of the manuscript. The authors thank all children and families who participated in this study.

